# BrachyAtlas: Patient-Specific Virtual Planning for Intracavitary Cesium-131 Brachytherapy with Tile Placement, Dose Calculation, and Modeled Neuroanatomical Exposure

**DOI:** 10.64898/2026.09.06.26362396

**Authors:** Siyar Bahadir, Griffin Thomas, Faina Ablyazova, Sandra Leskinen, Marcio Y. Ferreira, Todd A. Goldstein, Netanel Ben-Shalom, A.Gabriella Wernicke

## Abstract

**Background:** Brachytherapy delivers localized radiation from sources placed within or adjacent to tissue at risk. In brain tumor surgery, intracavitary implantation can begin at resection and concentrate dose along the cavity wall, where many recurrences arise. Current preoperative GammaTile tools estimate tile requirements but not patient-specific placement, resulting dose, or adjacent anatomy. We developed an integrated software framework for patient-specific virtual GammaTile planning.

**Methods:** The framework incorporates AI-assisted tumor and cavity segmentation with user approval, converts accepted masks into patient-derived surfaces, supports virtual tile placement, calculates lifetime Cs-131 dose using TG-43, maps dose distributions to HCP-MMP cortical parcels and normative HCP-1065 white-matter bundles, and supports AI-assisted synthesis of the resulting complex anatomical and connectomic output for clinician review. We retrospectively applied it to three patients with glioblastoma. Tile requirements were compared with the GammaTile Cavity Surface Area Calculator. For Patient 1, calculated 60- and 80-Gy volumes were compared with the clinical plan.

**Results:** Preoperative layouts required 6.5, 7, and 4 tile equivalents for Patients 1–3; calculator estimates were higher by 1.5, 4, and 1 tiles. Postoperative differences narrowed to 1, 1, and 0 tiles. The plans generated patient-specific lifetime dose distributions at 60, 80, and ≥120 Gy. In Patient 1, calculated and clinical 60- and 80-Gy volumes were similar despite incomplete spatial overlap. Atlas mapping revealed distinct cortical and white-matter exposure patterns across cases beyond lobar description.

**Conclusions:** Patient-specific virtual GammaTile planning can connect anticipated tile placement with calculated dose and adjacent neuroanatomy. These cases demonstrate feasibility and establish a framework for prospective validation and future comparison of candidate implant arrangements.

## 1. Background

Brachytherapy delivers radiation from sources placed within or immediately beside tissue at risk, producing a high local dose with a steep falloff over distance. In intracranial tumors, this principle is especially relevant because many recurrences arise within or immediately beside the resection bed [1–3]. Placing sources along the cavity wall at surgery can begin localized treatment without waiting for postoperative healing and external-beam radiotherapy.

Intracavitary brachytherapy has been implemented with permanent seeds positioned along the operative bed, where source position, orientation, and spacing determine both cavity coverage and dose to adjacent brain [4–6]. GammaTile is a standardized implementation: four Cs-131 seeds are embedded in a 20 × 20 mm collagen carrier that maintains source spacing and separates the seeds from the tissue surface. The short Cs-131 half-life provides a high initial dose rate, while the carrier constrains local geometry. The device has been described in recurrent meningioma, brain metastasis, and glioma series [7–10]. Prospective evaluation includes the randomized phase III ROADS trial in resected brain metastases and the phase IV GESTALT trial in newly diagnosed glioblastoma [11,12], increasing the importance of reproducible planning before implantation.

Although GammaTile standardizes source spacing after placement, patient-specific cavity geometry remains uncertain before and during surgery and continues to change afterward. Reported source migration averages approximately 1.8 mm and reaches 7.3 mm, GammaTile displacement approaches 6 mm as the cavity changes, and cavity volume contracts with an effective half-life of roughly three months [13–15]. Delivered D90 has ranged from 31.7 to 98.7 Gy against a 60-Gy prescription [6]. Current preoperative tools therefore focus primarily on inventory. The GammaTile Cavity Surface Area Calculator uses three cardinal dimensions, an ellipsoidal representation, expected contraction, and a deduction for untreated surface [16], whereas newer ordering tables use maximum tumor dimension and histology [17,18]. In 15 cavities, Chaswal and colleagues found frequent overestimation by both dimension- and contour-based workflows, particularly for cavities larger than 30 cmZ, while the contour-based method sometimes underestimated requirements [19]. These approaches do not show where individual tiles could fit on an irregular anticipated cavity, the dose field produced by a candidate arrangement, or which nearby cortical parcels and white-matter pathways that field would intersect. Individual tile positions are still selected after the cavity becomes visible, with postimplant imaging used to reconstruct dose [5,6,9,13–15].

Several studies have refined preoperative planning. Allen and Ferreira represented full and half GammaTiles as rigid applicators in BrachyVision and manually translated and rotated each applicator to create a presurgical plan with real-time dose estimates [20]. Huang et al used a patient-specific three-dimensional printed surrogate before the first reported GammaTile spine implantation [21]. Earlier planar permanent-seed methods established rapid intraoperative optimization and source-strength calculations [22,23], and comparative dosimetric studies placed GammaTile dose distributions in the context of stereotactic radiotherapy [4]. These reports establish the feasibility of earlier dose estimation, although published intracranial approaches still require manual placement of rigid tile representations or reduce the anticipated cavity to a small set of dimensions.

A second line of work has made white-matter anatomy increasingly visible to radiation planning. Diffusion tractography has been incorporated into Gamma Knife, stereotactic, and intensity-modulated planning for the optic radiations, pyramidal tracts, and motor pathways [24–29].

Automated atlas-based tract mapping extends this direction toward reproducible pathway-aware planning [30], and the COG-SRS study demonstrates that anatomy-derived structures can carry prospective dose constraints [31]. Longitudinal imaging studies also show that radiation effects vary among white-matter bundles and along tract axes, including perisylvian, callosal, intrahemispheric, and limbic pathways [32–37]. These findings support describing dose at the bundle level rather than relying only on whole-brain or lobar summaries.

What remains missing is a shared patient-specific representation linking anticipated resection geometry, possible tile and seed locations, calculated dose, and adjacent neuroanatomy. A tile count alone does not provide source coordinates, and these components are generally resolved at different times or in separate systems.

We therefore developed BrachyAtlas, an AI-assisted, human-supervised browser-based framework integrating tumor/cavity segmentation, patient-specific surface reconstruction, interactive virtual tile placement, TG-43 dose calculation, atlas-based anatomical exposure analysis, and AI-assisted synthesis of the complex anatomical and connectomic output for clinician review. We applied it to three retrospective glioblastoma cases, compared virtual tile requirements with the GammaTile calculator, benchmarked the dose calculation, evaluated the available clinical postimplant plan, and summarized parcel- and tract-level exposure. The workflow extends atlas-based mapping used in external-beam stereotactic radiotherapy [30] and the anatomy-guided planning paradigm of COG-SRS [31].

## 2. Methods

### 2.1 Study design, cases, and imaging

This retrospective proof-of-concept study included three patients with glioblastoma treated using intracavitary Cs-131 collagen tiles: a left parieto-temporal lesion (Patient 1), right frontal lesion (Patient 2), and diffuse right fronto-temporo-parietal lesion (Patient 3). Preoperative and postoperative volumetric MRI were available for all patients. Enhancing disease was delineated on contrast-enhanced T1-weighted imaging; the nonenhancing component resected and tiled in Patient 3 was delineated on FLAIR. Patient 1 also had a clinical postimplant CT and dose export (Supplementary Table E4). This retrospective study was approved by the institutional review board at Northwell Health (IRB No. 26-0214-NH).

### 2.2 Segmentation, surface construction, and planning map

Preoperative tumor and postoperative cavity masks were exported in NIfTI or NRRD format. Preoperative masks provided patient-derived representations of projected resection surfaces, whereas postoperative masks represented observed cavities for retrospective comparison.

BrachyAtlas incorporates AI-assisted segmentation through Vox2Tell, using the VoxTell free-text-promptable three-dimensional segmentation framework [38]; users may alternatively draw a mask manually, import an external mask, or edit an automated result. Segmentation performance was not evaluated, and all analyses used the user-approved mask as input.

Each mask was converted to a three-dimensional mesh and divided into brain-facing regions eligible for coverage and excluded regions facing skull or another untreated surface. The curved mesh was parameterized by axial level and meridian and opened along a selected vertical meridian to create a two-dimensional planning map linked pointwise to the original three-dimensional coordinates. Tiles were positioned on this map, while spacing, orientation, seed coordinates, and dose calculation remained defined on the patient-derived surface (Figure 2; Supplementary Figures E1 and E2).

### 2.3 Spatial registration

Patient images were registered to Montreal Neurological Institute (MNI) space using centering, rigid and similarity transformations optimized with mutual information, and symmetric diffeomorphic normalization optimized with cross-correlation [39]. Preoperative and postoperative images were additionally aligned by rigid registration. Registration was inspected visually at ventricular, cortical, and lesion-adjacent boundaries (Figure 1C,D; Figure 5D; Supplementary Figures E1D and E2D). Nearest-neighbor interpolation was used for categorical masks and continuous interpolation for dose volumes.

**Figure 1.**
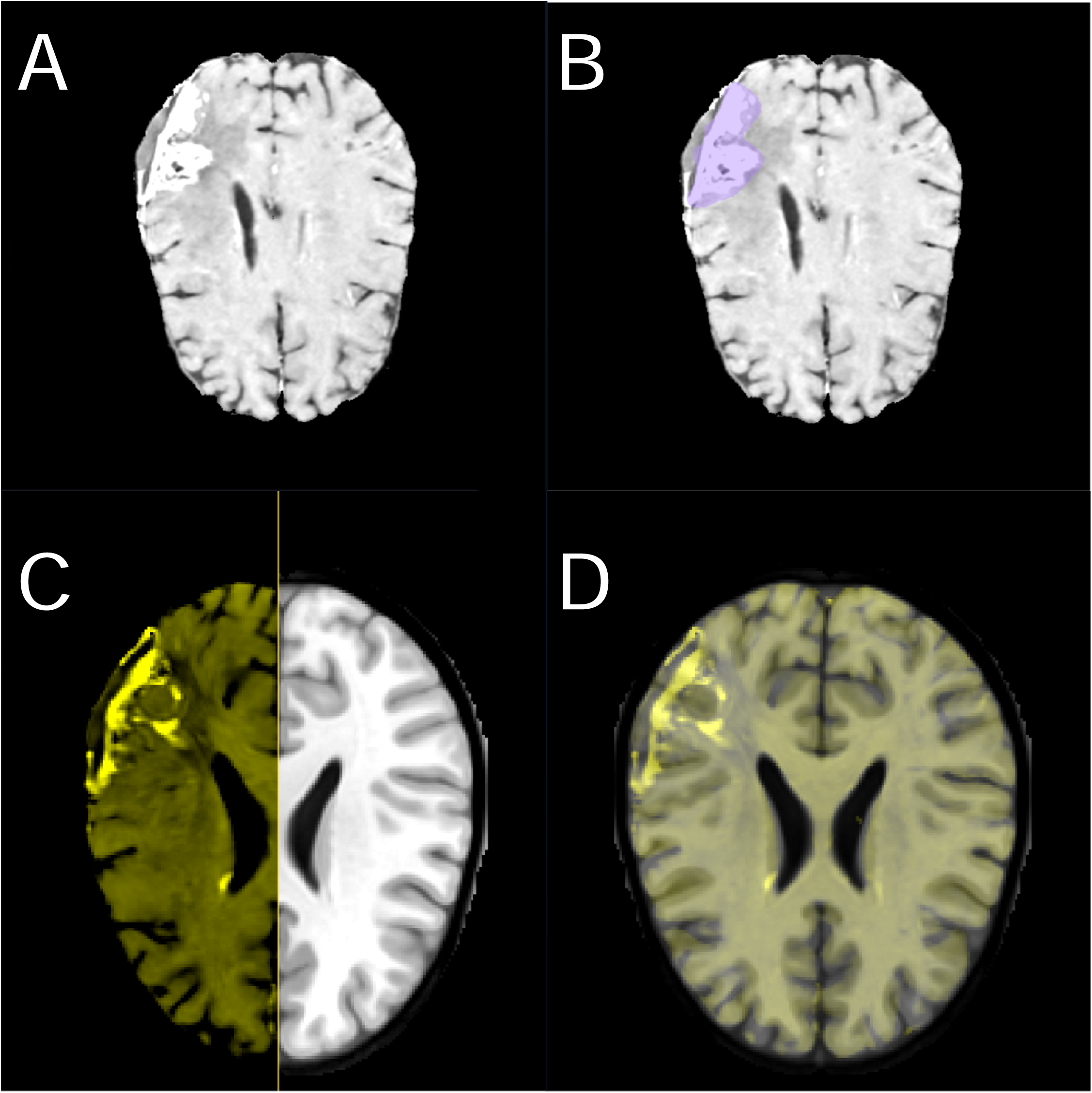
Patient 1 preoperative target segmentation and registration to standard space. (A) Axial preoperative MRI showing the observed left parieto-temporal lesion. (B) The same slice with the patient-derived target segmentation in violet; this mask supplied the projected-resection surface representation. (C) Split-screen comparison of the registered patient image in yellow and the MNI template in gray. (D) Blended patient-template overlay used to inspect ventricular, cortical, and lesion-adjacent alignment.

### 2.4 Virtual tiles and Cs-131 dose calculation

Each virtual tile represented a 20 × 20 mm collagen carrier containing four Cs-131 seeds in a 2 × 2 arrangement at 1-cm center-to-center spacing [5,7,9]. The seed plane was positioned approximately 3 mm inside the cavity void. Placement followed local surface curvature and excluded untreated regions. One tile equivalent was defined as four active seed positions; preoperative surfaces were uniformly contracted by 20% before placement and dose calculation. Each active seed was modeled as an IsoRay CS-1 Rev 2 line source. Dose rate was calculated using the AAPM TG-43 geometry, radial-dose, and anisotropy functions with the source-specific air-kerma strength and dose-rate constant [40–44]. Contributions were summed on a three-dimensional grid and integrated to infinity using the 9.7-day Cs-131 half-life. The calculation represented dose to water in water and did not model tissue or material heterogeneity [45]. The Patient 1 clinical comparison used 3.03 U per seed. Calculated 60-, 80-, and 120-Gy contours represented the prescription comparator, an additional available clinical comparator, and the high-dose region surrounding the sources, respectively (Figures 3 and 5).

### 2.5 GammaTile calculator comparison

Virtual requirements were compared with the GammaTile Cavity Surface Area Calculator [16,19], which estimates a three-axis ellipsoid, deducts expected contraction and untreated surface, divides the remaining area by 4 cmZ, and rounds upward. The implementation reproduced calculator outputs within 1%. Both preoperative methods used 20% contraction; postoperative comparisons used 0%. The untreated-surface fraction was specified per case (Table 1).

**Table 1.** GammaTile calculator estimates versus patient-specific virtual layouts before and after resection.

| Case | Phase | AP × ML × SI<br>(mm) | Calculator<br>ellipsoid<br>surface<br>(cm<sup>2</sup>) | Contraction<br>/ untreated | Area<br>needing<br>tiles (cm<sup>2</sup>) | GammaT<br>ile<br>calculato<br>r<br>estimate | Virtual<br>seeds (tile<br>equivalents) | Coverage<br>(mm<sup>2</sup>) |
| --- | --- | --- | --- | --- | --- | --- | --- | --- |
| 1 | Pre | 57.88 × 24.92 ×<br>63.82 | 75.1 | 20% / 50% | 30.0 | 8 | 26 (6.5) | 2014 |
| 1 | Post | 56.70 × 17.01 ×<br>36.74 | 41.7 | 0% / 50% | 20.9 | 6 | 20 (5) | 1824 |
| 2 | Pre | 48.00 × 52.09 ×<br>34.85 | 63.4 | 20% / 20% | 40.6 | 11 | 28 (7) | 2414 |
| 2 | Post | 23.34 × 49.54 ×<br>33.60 | 39.1 | 0% / 20% | 31.3 | 8 | 28 (7) | 2029 |
| 3 | Pre | 33.16 × 34.82 ×<br>19.46 | 26.3 | 20% / 20% | 16.8 | 5 | 16 (4) | 1067 |
| 3 | Post | 29.74 × 24.05 ×<br>14.65 | 16.4 | 0% / 20% | 13.1 | 4 | 16 (4) | 995 |

Note. Pre = projected preoperative resection surface; Post = observed postoperative cavity; AP = anterior–posterior; ML = medial–lateral; SI = superior–inferior. Area needing tiles is the calculator ellipsoid surface after contraction and untreated-surface deductions. One tile equivalent equals four virtual seed positions; fractional equivalents are computational summaries rather than fractional manufactured inventory.

### 2.6 Dose benchmarking and clinical-plan comparison

The dose engine was benchmarked at the 1-cm reference point against the published Carleton Laboratory for Radiotherapy Physics TG-43 calculation [43]. Patient 1 enabled an additional comparison between the saved 20-seed postoperative virtual plan and clinical 60- and 80-Gy isodose surfaces. Clinical surfaces were converted to binary NIfTI masks on their original 0.518 × 0.518 × 1.00 mm grid without smoothing or resampling. Virtual dose was recomputed on a 1.5-mm isotropic grid with a 45-mm margin, sampled by trilinear interpolation at clinical-grid voxel centers, and thresholded at matching levels (Figure 5).

Volumetric agreement included field volumes, shared and unique volumes, volume ratio, Dice and Jaccard indices, sensitivity, positive predictive value, and centroid distance. Clinical and calculated binary volumes were also transformed to MNI space using the saved forward diffeomorphic transform. A normative streamline was counted when at least one sampled point entered a volume; set-level and bundle-level agreement included overlap metrics and correlations of within-bundle percentages for bundles containing at least 10 streamlines. Analyses were descriptive because only one clinical comparator was available (Table 2; Supplementary Table E5). Additional implementation details are provided in Supplementary Methods.

**Table 2.** Selected agreement measures for Patient 1 calculated and clinical isodose volumes.

| **Metric** | **80 Gy** | **60 Gy** |
| --- | --- | --- |
| Volumetric agreement |  |  |
| Clinical volume (mL) | 20.89 | 31.63 |
| Calculated volume (mL) | 21.33 | 31.89 |
| Dice similarity | 0.7393 | 0.8021 |
| Centroid offset (mm) | 3.48 | 3.56 |
| Normative HCP-1065 streamline-set and bundle agreement |  |  |
| Clinical streamlines captured | 98.6% | 100.0% |
| Streamline-set Dice | 0.8478 | 0.7406 |
| Bundle Spearman $\rho$ (bundles compared) | 0.9604 (14) | 0.7887 (17) |

Note. The calculation used 3.03 U per seed. Streamline metrics use HCP-1065 trajectories; clinical streamlines captured is the fraction of the clinical set recovered by the calculation. Bundle Spearman $\rho$ compares percentages within eligible bundles containing at least 10 streamlines. Full results appear in Supplementary Table E5.

### 2.7 Atlas-based cortical and white-matter analysis

Registered dose fields were evaluated relative to the 360-region HCP-MMP parcellation [46] and the population-averaged HCP-1065 tractogram, comprising 10,403 streamlines in 106 named bundles derived from generalized q-sampling imaging [47–49]. These normative trajectories are modeling units rather than patient-specific fibers or biological counting units and remain subject to tractography and interindividual limitations [50,51].

Parcels were ranked within each case by maximum sampled dose. Each streamline was assigned by the maximum dose sampled along its trajectory; ≥120 Gy was cumulative, while lower categories were bounded by the next higher threshold. Projected-resection and calculated-dose intersections were queried separately and could overlap: the former counted any trajectory entering the projected-resection mask, whereas the latter assigned the same trajectories by sampled dose. Bundle outputs were counts and within-bundle percentages after excluding bundles with fewer than 10 streamlines. These values describe modeled anatomical exposure rather than biological injury or clinical outcome (Table 3; Supplementary Tables E1–E3).

**Table 3.**
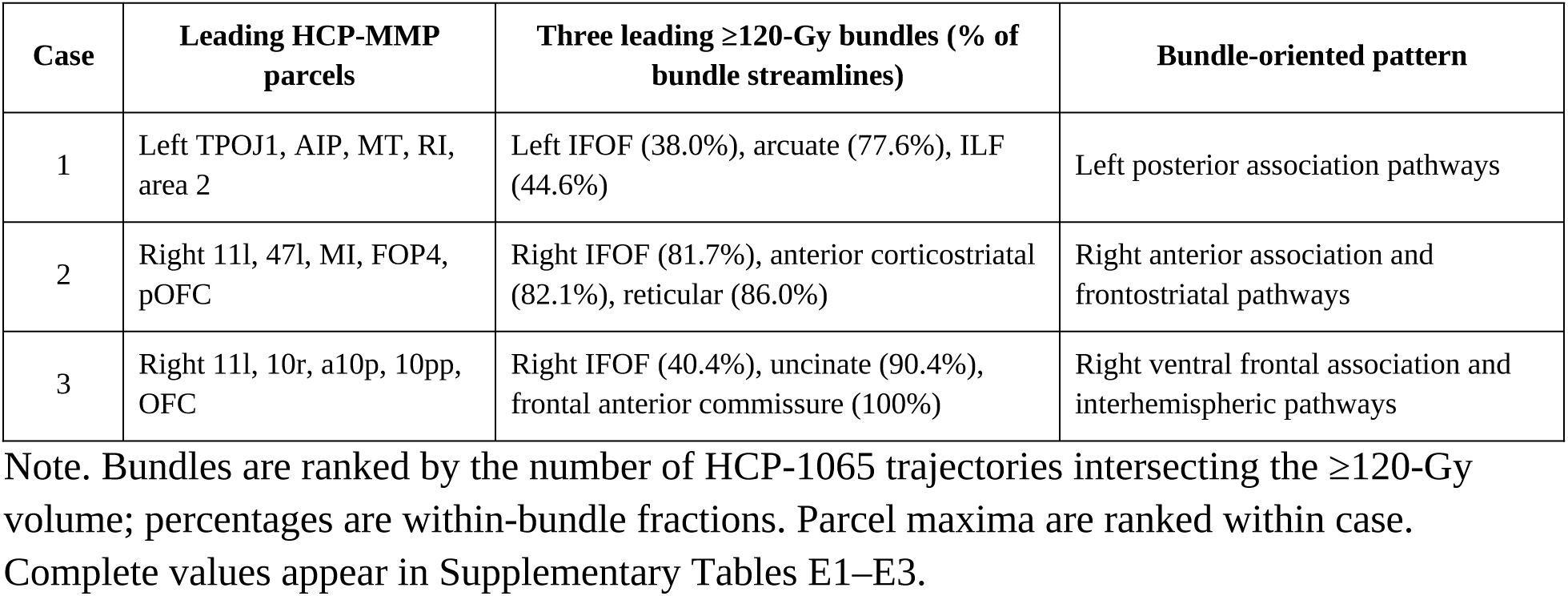
Leading cortical parcels and bundles intersecting the ≥120-Gy calculated dose volume.

### 2.8 Software implementation

BrachyAtlas is a browser-based application backed by a Python analysis pipeline. Imaging, segmentations, processing jobs, and plans are stored in a Patient-Scan-Job-Plan hierarchy, allowing arrangements to be reopened and recalculated without repeating completed steps. Interactive placement updates the corresponding three-dimensional footprints, seed coordinates, and tile totals. Structured parcel-, bundle-, and dose-level outputs can then be passed to an AI-assisted module for synthesis of the complex anatomical and connectomic output for clinician review. The AI-assisted synthesis does not generate or modify the dose distribution, registration transform, atlas assignments, or quantitative exposure metrics. Dose recomputation completes in under one minute on standard hardware, while registration and atlas sampling are reused across plans from the same scan. Extended workflow and implementation details are provided in Supplementary Methods. BrachyAtlas also incorporates a context-aware AI assistant that can provide page-specific guidance and respond to questions throughout the analysis workflow.

The application is a research prototype that has not undergone clinical commissioning, regulatory review, or treatment-planning quality assurance. All reported analyses were retrospective and did not inform treatment.

## 3. Results

### 3.1 Integrated planning representations

BrachyAtlas generated a linked surface, planning map, virtual implant, dose volume, structured anatomical/connectomic output, and reviewable AI-assisted synthesis for all three cases (Figures 1-4; Supplementary Figures E1 and E2). Patient 1 additionally enabled the clinical-plan comparison shown in Figure 5.

### 3.2 Tile requirements: GammaTile calculator versus virtual layout

For every preoperative case, the GammaTile calculator produced a larger estimate than the virtual layout: 1.5 tile equivalents for Patient 1, 4 for Patient 2, and 1 for Patient 3. Postoperative differences were 1, 1, and 0, respectively; Patient 3 showed exact agreement. The Patient 2 postoperative layout contained 28 virtual seed positions, equal to seven tile equivalents (Table 1).

Preoperative rows compare calculator estimates with virtual tiles placed on projected resection surfaces; postoperative rows use the observed cavities, bringing both methods closer to the final implant geometry (Table 1; Figure 2; Supplementary Figures E1 and E2).

**Figure 2.**
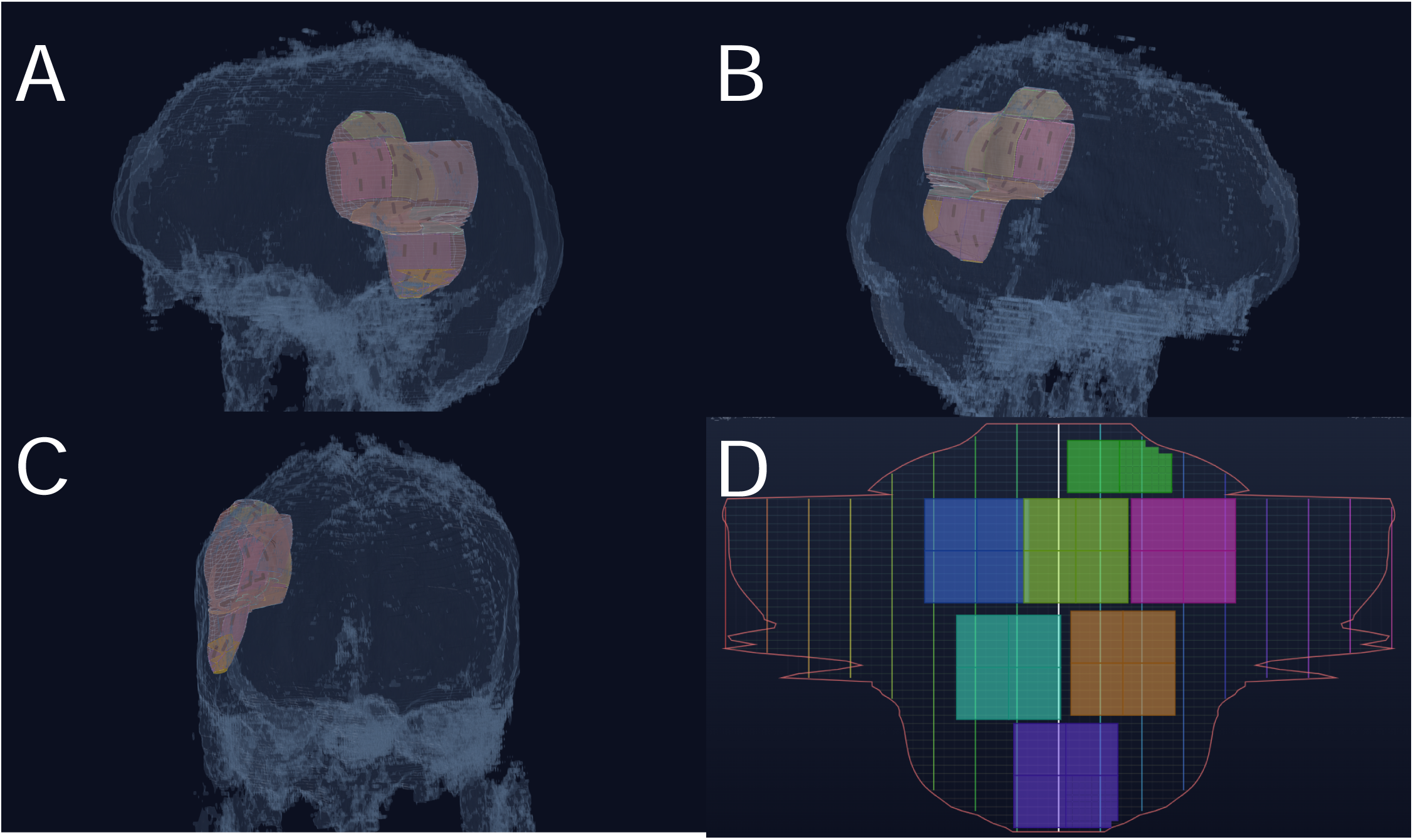
Virtual tile placement on the patient-derived representation of Patient 1’s projected preoperative resection surface. (A) First three-dimensional oblique view of the surface representation and colored virtual tile carriers. (B) Opposite oblique view showing coverage across the superior and lateral surface. (C) Coronal view showing the superior-inferior distribution of the virtual tiles. (D) Opened planning map; the red contour marks the modeled surface boundary, the grid retains correspondence to the three-dimensional surface representation, and each colored rectangle represents a virtual tile footprint.

### 3.3 Dose benchmarking and agreement with the clinical plan

For Patient 1, the virtual seed pattern visually corresponded to seeds on postimplant imaging, and calculated 60- and 80-Gy bands overlapped the clinical boundaries in representative sections (Figure 5B,E,F).

Calculated and clinical field sizes were closely matched: 21.33 versus 20.89 mL at 80 Gy and 31.89 versus 31.63 mL at 60 Gy. Dice was 0.7393 and 0.8021, with centroid offsets of 3.48 and 3.56 mm, respectively (Figure 5F; Table 2; Supplementary Table E5).

The calculated volume recovered 98.6% of the clinical streamline set at 80 Gy and 100.0% at 60 Gy. Streamline-set Dice was 0.8478 and 0.7406, and bundle-level percentages remained strongly associated (Spearman ρ, 0.9604 and 0.7887; Figure 5G; Table 2).

The largest bundle-level discrepancies were the left arcuate fasciculus at 80 Gy (69.9% calculated versus 29.1% clinical) and the left inferior longitudinal and inferior fronto-occipital fasciculi at 60 Gy (45.0% versus 1.8% and 38.7% versus 10.5%, respectively; Figure 5G).

### 3.4 Case-specific bundle exposure patterns

At ≥120 Gy, the leading bundles were the left inferior fronto-occipital, arcuate, and inferior longitudinal fasciculi in Patient 1; the right inferior fronto-occipital, anterior corticostriatal, and reticular bundles in Patient 2; and the right inferior fronto-occipital and uncinate fasciculi and frontal anterior commissure in Patient 3. The two broadly right-frontal cases therefore differed in their anterior frontostriatal versus ventral frontal and interhemispheric patterns (Figure 4; Supplementary Figures E1 and E2; Table 3). Complete rankings appear in Supplementary Tables E1–E3.

## 4. Discussion

This study links patient-specific virtual GammaTile planning to calculated dose and atlas-based anatomical interpretation. The principal findings were higher preoperative estimates from the GammaTile calculator than from the virtual layouts, close Patient 1 isodose volumes despite residual spatial disagreement, and bundle patterns that distinguished anatomically different cases (Tables 1–3).

Clinical trials provide context. In the 2026 ROADS phase III trial of resected brain metastases, 12-month surgical-bed recurrence was 1.3% with Cs-131 tile-based radiation versus 15.4% with postoperative stereotactic radiation, with comparable grade ≥3 adverse events [11]. The phase IV GESTALT trial evaluates GammaTile as an intraoperative boost with standard chemoradiation for newly diagnosed glioblastoma [12]. These trials do not validate BrachyAtlas but make reproducible patient-specific planning increasingly relevant.

### 4.1 Beyond tile inventory: the modeled cavity surface

Calculator and ordering tools support preoperative inventory [16–19] but do not specify how square tiles can be distributed over a surface whose circumference, curvature, and untreated regions vary. The opened map makes these features explicit and links each virtual footprint to patient coordinates (Figure 2; Supplementary Figures E1 and E2).

The largest differences in Table 1 occurred when both methods began with preoperative tumor geometry; differences narrowed after the cavity became observable. Chaswal and colleagues similarly reported frequent overestimation, particularly for cavities larger than 30 cmZ [19]. The calculator begins with an ellipsoid and fixed deductions, whereas the virtual method places tile representations on selected portions of an irregular surface. Underestimation risks an intraoperative shortfall of a manufactured, calibrated source, while overestimation adds radioactive-material handling and storage burden.

The comparison does not establish that the virtual preoperative count is more accurate. The future cavity may differ from the tumor-derived surface because of operative strategy, collapse, hemostatic material, untreated surface, contraction, and source displacement [13–15].

Prospective validation should compare the preoperative projection, intraoperative cavity, implanted positions, and serial postimplant imaging.

### 4.2 From seed coordinates to a preoperative dose forecast

Once a virtual arrangement supplies source coordinates and strength, TG-43 dose can be calculated before surgery [40–44] (Figure 3). Patient 1 showed why matched isodose volumes do not establish spatial agreement: similar field sizes coexisted with incomplete overlap (Figure 5F; Table 2).

**Figure 3.**
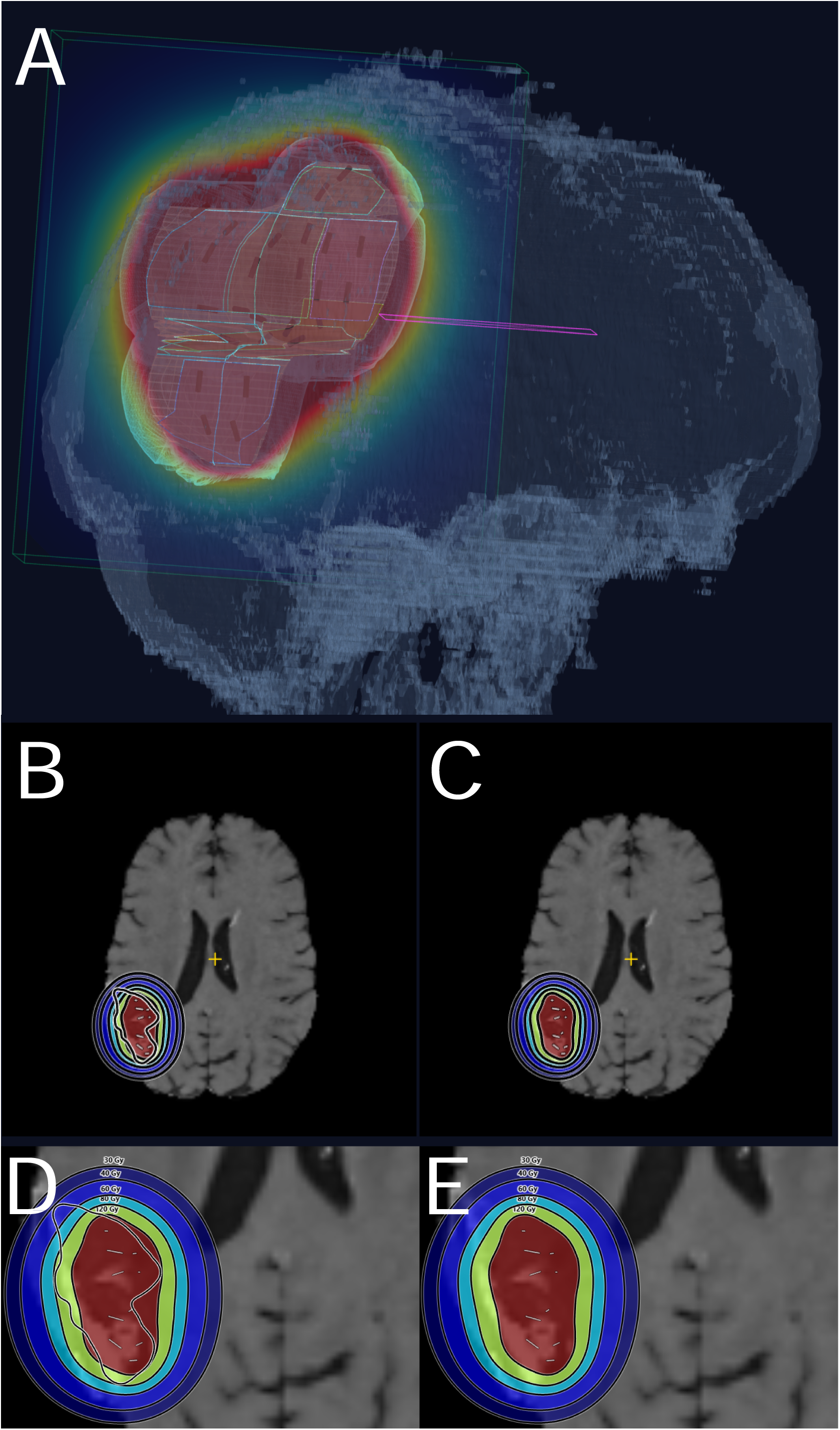
Preoperative TG-43 lifetime dose calculation for the Patient 1 virtual implant. (A) Three-dimensional calculated dose field surrounding the patient-derived representation of the tiled projected-resection surface; short dark marks represent virtual Cs-131 seeds. (B) Axial overview on the preoperative MRI with the 30-, 40-, 60-, 80-, and 120-Gy calculated dose bands and the target boundary. (C) The same axial level with the target boundary suppressed. (D) Magnified view of panel B. (E) Magnified view of panel C. Red denotes the highest displayed dose band, followed outward by green, cyan, and blue bands.

The approximately 3.5-mm centroid offset was predominantly axial and was measured in the native clinical grid, independent of patient-to-MNI registration. Because the comparison used the observed postoperative cavity without contraction, the offset may instead reflect virtual-versus- implanted seed positions, source localization, cavity delineation, or dose reconstruction (Figure 5B,F).

Earlier work manually positioned rigid GammaTile applicators in a commercial planning system [20] or used a three-dimensional surrogate for spine implantation [21]. Atlas-based tract dosimetry has also been demonstrated for external-beam stereotactic radiotherapy [30], and COG-SRS showed that anatomy-derived structures can carry prospective dose constraints [31].

BrachyAtlas connects these previously separate ideas by carrying virtual intracavitary tile coordinates into TG-43 dose calculation and HCP-MMP cortical and normative bundle mapping (Figures 2–4).

The streamline comparison adds an anatomical measure of projected-versus-realized field agreement. In Patient 1, the clinical sets were almost entirely recovered, while calculated-only trajectories concentrated in long association bundles near the field periphery (Figure 5G; Table 2). Because one sampled point was sufficient to count a trajectory, these fractions emphasize boundary involvement rather than irradiated path length.

This remains a single-case validation. Broader evaluation should include multiple clinical dose exports, quantitative seed matching, serial cavity geometry, voxelwise comparisons when full dose grids are available, and cavity-shell dose-volume metrics. Model-based calculations could then address tissue and implant heterogeneity beyond TG-43 dose to water [5,6,13–15,45].

### 4.3 Projected resection and calculated dose in one atlas representation

Reporting projected resection and calculated dose in one atlas places their relationship to each bundle side by side (Figure 4; Supplementary Figures E1 and E2; Supplementary Tables E1–E3). Because the queries may overlap, they describe the combined operative and radiation environment without assigning a postoperative change to either mechanism. Causal separation would require segment-level labels, longitudinal imaging and outcomes, and consideration of progression, edema, ischemia, medication, and adjuvant treatment.

**Figure 4.**
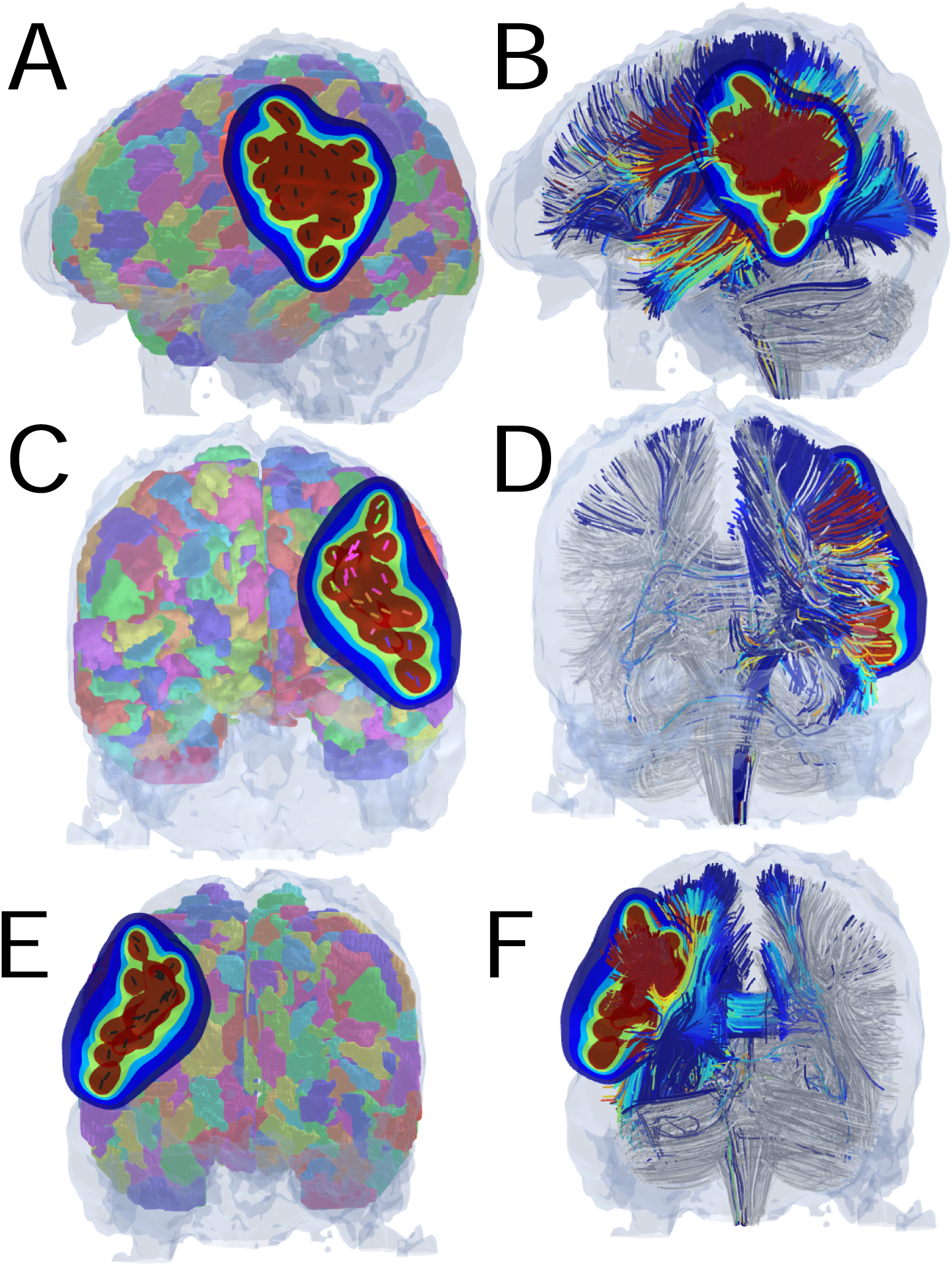
Patient 1 preoperative dose mapping to cortical parcels and normative white-matter bundles. (A) Lateral view of HCP-MMP cortical parcels with calculated dose bands overlaid. (B) Corresponding lateral view of the normative HCP-1065 tractogram. (C) Anterior cortical view. (D) Corresponding anterior tractogram view. (E) Posterior cortical view. (F) Corresponding posterior tractogram view. Atlas streamlines are colored by the maximum sampled dose along their trajectories; gray streamlines lie below the displayed dose bands.

### 4.4 The insufficiency of lobar description

Although Patients 2 and 3 were both broadly right frontal, Patient 2 showed an anterior association and frontostriatal pattern, whereas Patient 3 showed a more ventral frontal and interhemispheric pattern; Patient 1 showed a separate left posterior association pattern (Figure 4; Supplementary Figures E1 and E2; Table 3).

Radiation-associated white-matter changes vary among bundles and along tract axes [32–34]. Perisylvian injury has been associated with language decline [35], callosal and intrahemispheric injury with attention and processing-speed decline [36], and limbic injury with memory performance [37]; voxel-based and cortical studies likewise indicate regional heterogeneity [52–54]. These findings support anatomically resolved dose descriptions but do not provide transferable tolerance values for short-range permanent Cs-131. The present fractions and parcel rankings therefore remain geometric rather than predictive of symptoms or outcomes.

### 4.5 Candidate arrangements and changing cavity geometry

BrachyAtlas can compare candidate tile arrangements on the same surface. External-beam studies show that functional cortex, tracts, and hippocampi can be incorporated into planning while maintaining target goals [24–29,31,54–57]. GammaTile offers different control variables —tile position, orientation, spacing, and omission from selected regions—which the opened map makes computationally accessible. The present cases establish the representation needed for such comparisons but do not show that an alternative arrangement improves function.

A further extension could couple virtual implants to patient-specific finite-element models of postoperative contraction. Propagating tiles through a predicted deformation field and integrating recalculated dose-rate distributions during Cs-131 decay would estimate cumulative dose under changing cavity geometry rather than assuming a static configuration.

The public BrachyAtlas release allows independent users to inspect assumptions, reproduce the output structure, and test alternative arrangements or anatomical analyses on institutionally approved data. Availability supports reproducibility and method development but does not constitute commissioning, regulatory clearance, or validation for clinical treatment planning. The AI components occupy bounded roles at the input and interpretation ends of the workflow. AI-assisted segmentation can accelerate creation of an initial patient-specific mask while preserving clinician approval, and AI-assisted synthesis can organize high-dimensional parcel-, bundle-, and dose-level outputs for clinician review. The intervening geometry, tile coordinates, TG-43 dosimetry, registration, and atlas sampling remain explicit computational outputs. The synthesis is therefore an interface to the calculated results rather than a substitute for radiation- physics or clinical judgment, and its performance as an interpretive aid was not independently evaluated in this study. This context-aware assistance may further reduce the technical burden of navigating complex imaging, dosimetric, and connectomic analyses while maintaining clinician oversight.

### 4.6 Limitations

The cohort contains three retrospective cases, and only Patient 1 had a clinical comparator (Supplementary Table E4). Preoperative surfaces were based on tumor geometry and cannot reproduce every operative decision or later cavity shape. Patient 3 target definition was informed by postoperative review and therefore tests geometric workflow rather than prospective delineation. The models applied uniform 20% contraction but not time-dependent deformation or source displacement [13–15]. Clinical comparison used binary 60- and 80-Gy masks rather than a complete dose grid, preventing dose-difference or gamma analysis; seed correspondence remained visual (Figure 5; Supplementary Table E5). Calculations used a 1.5-mm grid and TG-43 without tissue or material heterogeneity [40,45].

**Figure 5.**
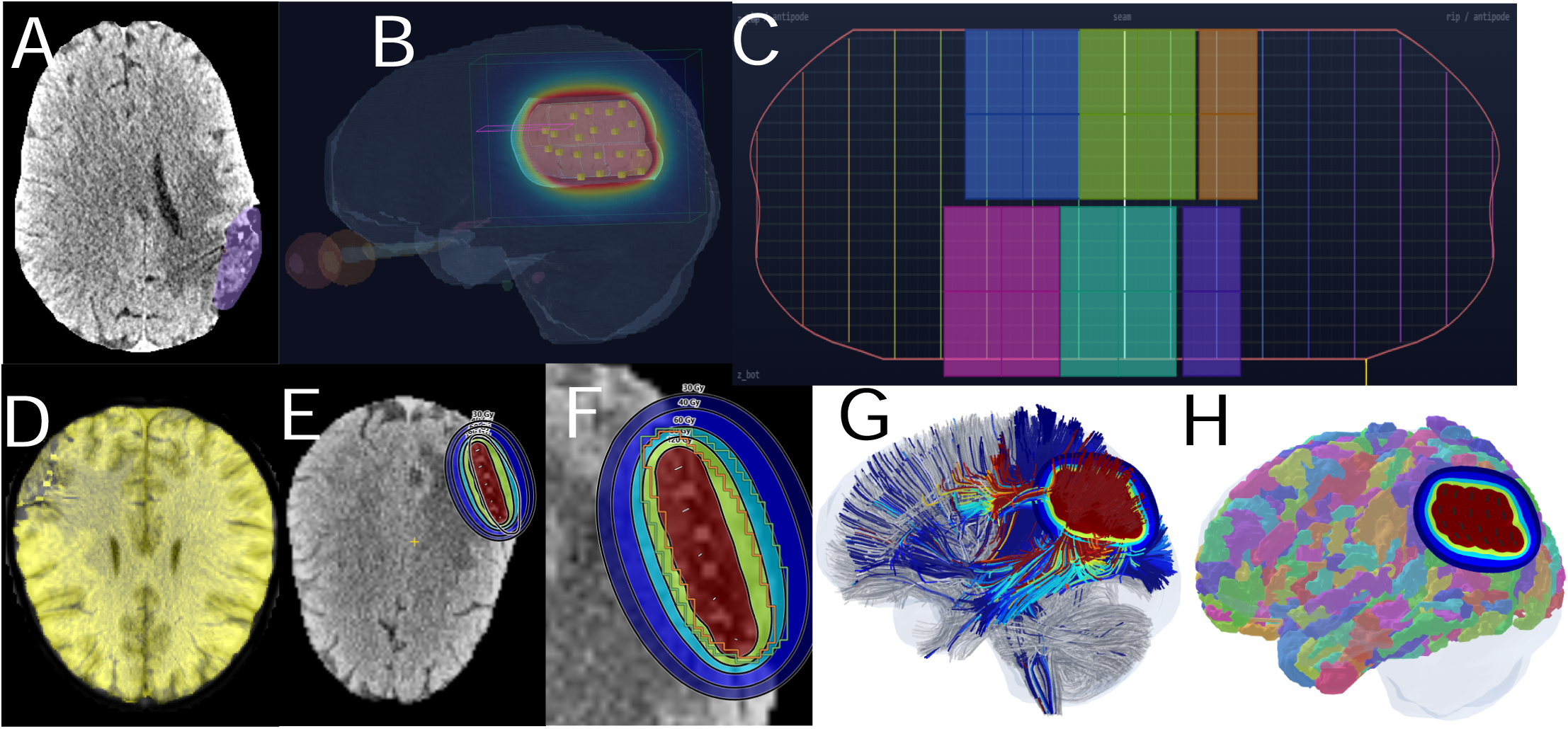
Patient 1 postoperative single-case comparison between the virtual and clinical implant representations. (A) Axial postoperative image with the observed cavity segmentation in violet. (B) Three-dimensional calculated dose field around the virtual tiles; dark source marks are virtual seeds and yellow cubes are seeds identified on postimplant imaging. (C) Opened planning map for the patient-derived postoperative cavity representation with virtual tile footprints. (D) Blended postoperative patient-to-MNI registration overlay. (E) Axial overview of the calculated lifetime dose bands. (F) Magnified comparison of the calculated bands with the clinical 60-Gy boundary in green and 80-Gy boundary in orange. (G) Normative HCP-1065 tractogram with atlas streamlines colored by their maximum sampled calculated dose. (H) HCP-MMP cortical parcels with the calculated dose bands overlaid. The clinical-comparison calculation used 3.03 U per seed, matching the clinical plan setting specified before agreement was evaluated.

Registration of a distorted brain to a standard atlas introduces uncertainty. Quality checks were visual, and lesion-aware normalization was not applied systematically [39,58,59]. HCP-MMP and HCP-1065 resources are normative and do not capture individual displacement; tractography has false-positive and false-negative limitations [50,51]. Single-point intersections and maximum sampled parcel dose are sensitive to boundary position and seed proximity. Later studies should prioritize dose-volume, surface-area, path-length, and segment-level measures.

Projected-resection and dose queries were not mutually exclusive, and no prospective neurological or neurocognitive assessments were available. BrachyAtlas remains an uncommissioned research prototype.

## 5. Conclusions

Virtual GammaTile planning can be linked to TG-43 dose calculation and atlas-based mapping. Across three retrospective cases, virtual tile requirements differed from GammaTile calculator estimates, and bundle summaries added anatomical detail beyond lobar location. The dose engine matched its reference calculation, and Patient 1 established the feasibility of clinical-plan comparison while underscoring the need for prospective spatial validation. Future work should test projected-cavity accuracy, validate additional implants, verify spatial correspondence across templates, and compare candidate arrangements under matched coverage and bundle- involvement requirements.

The accompanying release of BrachyAtlas provides a reusable implementation for reproducing and extending this research workflow.

## Data Availability

De-identified patient imaging is subject to institutional and privacy restrictions. Derived code, parameter files, and aggregate outputs will be made available with the BrachyAtlas software release, subject to repository licensing and institutional policy.

## Declaration of Generative AI and AI-assisted technologies in the writing process

During the preparation of this work, the authors used ChatGPT (OpenAI) for drafting support, literature research support, post-writing checks, and language editing, and Claude (Anthropic) for software-development and coding support. After using these tools, the authors reviewed and edited the content as needed and take full responsibility for the content of the publication.

## Declarations

### Ethics Statement

This retrospective study was approved by the Northwell Health Institutional Review Board (IRB No. 26-0214-NH).

### Author responsible for statistical analyses

Siyar Bahadir, MD. Funding statement: No specific funding was received for this work.

## Acknowledgments

Generative artificial intelligence tools were used in the development of this work. ChatGPT (OpenAI) was used for drafting support, literature research support, post-writing checks, and language editing; Claude (Anthropic) was used primarily for coding and software-development support. AI-assisted outputs were reviewed, verified, and revised by the authors, who take responsibility for the final manuscript, analyses, and software.

