## Supplementary Methods, Figures E1-E2, and Tables E1-E5 for "BrachyAtlas: Patient-Specific Virtual Planning for Intracavitary Cesium-131 Brachytherapy with Tile Placement, Dose Calculation, and Modeled Neuroanatomical Exposure"

Supplementary Methods, Tables E1–E5, and Figures E1–E2

### Supplementary Methods

#### E1. Data organization and segmentation workflow

Imaging, segmentations, registration jobs, virtual layouts, and dose plans were organized in a Patient–Scan–Job–Plan hierarchy. This preserved the relationship between each source image, processing step, and derived plan without overwriting native coordinate information. The integrated Vox2Tell module can generate an initial tumor or resection-cavity mask from a free-text prompt; users may instead create a mask manually, import an externally prepared NIfTI or NRRD mask, or edit an automated result. The release-matched VoxTell checkpoint and inference configuration will be versioned with the software release.

#### E2. Surface unfolding, placement interface, and plan management

The mesh was represented on a regular grid indexed by axial level and meridian and opened along a selected vertical meridian. The planning view displayed changing circumference, curvature, eligible surface, and excluded regions while preserving a pointwise link to the original surface. Distance and tile spacing were evaluated using the original three-dimensional coordinates rather than distance on the opened map. Each placement updated the corresponding three-dimensional carrier footprint and active seed coordinates. Plans stored seed coordinates, tile equivalents, covered area, source strength, dose outputs, and anatomical summaries and could be reopened, modified, and recomputed without repeating upstream processing.

The user workflow consists of importing imaging; generating, importing, or editing a segmentation; constructing and classifying the surface; opening the planning map; placing and revising tiles; reviewing three-dimensional footprints and seed coordinates; calculating dose; registering outputs for cortical and white-matter sampling; reviewing AI-assisted synthesis of complex anatomical and connectomic output for clinician review; and saving or exporting the plan. Registration and atlas-sampling jobs are run once per scan and reused across plans. Clinical-plan comparison is implemented as an additional job that reads uploaded isodose masks and writes structured and tabular outputs. BrachyAtlas also incorporates a context-aware AI assistant that can provide page-specific guidance and respond to questions throughout the analysis workflow.

#### E3. Clinical comparator processing and agreement metrics

The available clinical export contained 60- and 80-Gy isodose surfaces, which were converted offline to binary NIfTI masks in the native patient frame and tagged with their dose levels. The masks remained on the original 0.518 × 0.518 × 1.00 mm lattice without smoothing or modification. The Patient 1 postoperative virtual plan contained 20 seeds at 3.03 U. Contributions were summed on a 1.5-mm isotropic grid spanning the seed bounding box plus a 45-mm margin. For each clinical threshold, calculated dose was sampled by trilinear interpolation at the center of every clinical-grid voxel and thresholded at the matching level, producing clinical and calculated binary volumes on the same lattice.

Volumetric measures included clinical, calculated, shared, and unique volumes; calculated-to-clinical volume ratio; Dice and Jaccard indices; sensitivity; positive predictive value; and Euclidean centroid distance. After nearest-neighbor transformation of both masks to MNI space, set-level streamline measures included shared and unique trajectories, recovery of the clinical set, count ratio, Dice, and Jaccard. For bundles containing at least 10 streamlines, agreement included the number and percentage intersected, Spearman and Pearson correlations, and overlap among the ten leading bundles. Analyses used NumPy, SciPy, NiBabel, and DIPY and were descriptive because only one clinical comparator was available.

#### E4. Atlas sampling and reporting definitions

Each HCP-MMP parcel was sampled from the registered calculated-dose field and ranked within case by its maximum value. Each HCP-1065 trajectory was assigned the maximum dose sampled along the streamline. The ≥120-Gy category was cumulative; lower categories were half-open bands bounded by the next threshold. Projected-resection intersection required at least one sampled point inside the projected mask, with no minimum path length. Resection and dose queries were evaluated separately but could include the same trajectory. Bundle summaries reported counts and within-bundle percentages after excluding bundles with fewer than 10 trajectories. These definitions support reproducible geometric reporting and do not represent axon counts, biological effect, or predicted neurological deficit.

### Supplementary Tables

Supplementary Tables E1–E3 report complete preoperative modeled-exposure outputs. Definitions and counting rules follow Section 2.7; projected-resection and calculated-dose queries may overlap. The ≥120-Gy category is cumulative, whereas the 60-Gy category contains maxima from 60 to <120 Gy. Cortical values are within-case HCP-MMP parcel maxima, and bundles containing fewer than 10 trajectories are excluded.

#### Table E1. Patient 1 — left parieto-temporal glioblastoma

*Left parieto-temporal glioblastoma, preoperative virtual plan.*

##### E1A. Bundles intersecting the projected resection

| **Bundle** | **Intersecting streamlines** | **Streamlines in bundle** | **% intersecting** |
| --- | --- | --- | --- |
| Arcuate Fasciculus L | 168 | 196 | 85.7% |
| Superior Longitudinal Fasciculus L 2 | 119 | 273 | 43.6% |
| Corpus Callosum Body | 80 | 400 | 20% |
| Parietal Aslant Tract L | 64 | 64 | 100% |
| Thalamic Radiation L Posterior | 61 | 111 | 55% |
| Corpus Callosum Tapetum | 59 | 290 | 20.3% |
| Inferior Fronto Occipital Fasciculus L | 52 | 447 | 11.6% |
| Superior Longitudinal Fasciculus L 3 | 48 | 53 | 90.6% |
| Corticostriatal Tract L Posterior | 39 | 118 | 33.1% |
| Corpus Callosum Forceps Major | 30 | 256 | 11.7% |
| Middle Longitudinal Fasciculus L | 23 | 42 | 54.8% |
| Inferior Longitudinal Fasciculus L | 19 | 278 | 6.8% |
| Thalamic Radiation L Superior | 17 | 183 | 9.3% |
| Acoustic Radiation L | 12 | 17 | 70.6% |
| Corticopontine Tract L Parietal | 11 | 76 | 14.5% |

##### E1B. Bundles by calculated-dose category — ≥120 Gy

| **Bundle** | **Streamlines in category** | **Streamlines in bundle** | **% in category** | **Peak dose (Gy)** |
| --- | --- | --- | --- | --- |
| Inferior Fronto Occipital Fasciculus L | 170 | 447 | 38% | 2036.8 |
| Arcuate Fasciculus L | 152 | 196 | 77.6% | 716 |
| Inferior Longitudinal Fasciculus L | 124 | 278 | 44.6% | 2300 |
| Superior Longitudinal Fasciculus L 2 | 94 | 273 | 34.4% | 1282.1 |
| Thalamic Radiation L Posterior | 79 | 111 | 71.2% | 2259.2 |
| Corpus Callosum Forceps Major | 77 | 256 | 30.1% | 1990.7 |
| Corpus Callosum Body | 61 | 400 | 15.2% | 1479.8 |
| Corticostriatal Tract L Posterior | 48 | 118 | 40.7% | 1776 |
| Corpus Callosum Tapetum | 46 | 290 | 15.9% | 523.8 |
| Middle Longitudinal Fasciculus L | 37 | 42 | 88.1% | 1369.9 |
| Parietal Aslant Tract L | 29 | 64 | 45.3% | 1564.1 |
| Superior Longitudinal Fasciculus L 3 | 28 | 53 | 52.8% | 896.2 |

##### E1C. Bundles by calculated-dose category — 60 to <120 Gy

| **Bundle** | **Streamlines in category** | **Streamlines in bundle** | **% in category** | **Peak dose (Gy)** |
| --- | --- | --- | --- | --- |
| Inferior Fronto Occipital Fasciculus L | 136 | 447 | 30.4% | 2036.8 |
| Corpus Callosum Tapetum | 97 | 290 | 33.4% | 523.8 |
| Optic Radiation L | 44 | 124 | 35.5% | 166.3 |
| Anterior Commissure Occipital | 40 | 109 | 36.7% | 157.6 |
| Corpus Callosum Forceps Major | 36 | 256 | 14.1% | 1990.7 |
| Corpus Callosum Body | 30 | 400 | 7.5% | 1479.8 |
| Inferior Longitudinal Fasciculus L | 28 | 278 | 10.1% | 2300 |
| Corticostriatal Tract L Posterior | 22 | 118 | 18.6% | 1776 |
| Superior Longitudinal Fasciculus L 2 | 20 | 273 | 7.3% | 1282.1 |
| Medial Lemniscus L | 16 | 161 | 9.9% | 116.9 |
| Extreme Capsule L | 10 | 41 | 24.4% | 1105.2 |
| Thalamic Radiation L Superior | 10 | 183 | 5.5% | 631.7 |

##### E1D. Cortical parcels by maximum sampled dose (HCP-MMP)

| **Parcel** | **Anatomical location** | **Max dose (Gy)** |
| --- | --- | --- |
| L_TPOJ1 | Inferior Parietal | 1297.6 |
| L_AIP | Intraparietal Sulcus | 1233.1 |
| L_MT | Occipital | 930.4 |
| L_RI | Temporal | 918.3 |
| L_2 | Sensorimotor | 801.9 |
| L_IP1 | Intraparietal Sulcus | 765.8 |
| L_MST | Occipital | 735.1 |

*Maximum calculated dose sampled within each HCP-MMP parcel; ranking is within-case only.*

#### Table E2. Patient 2 — right frontal glioblastoma

*Right frontal glioblastoma, preoperative virtual plan.*

##### E2A. Bundles intersecting the projected resection

| **Bundle** | **Intersecting streamlines** | **Streamlines in bundle** | **% intersecting** |
| --- | --- | --- | --- |
| Inferior Fronto Occipital Fasciculus R | 825 | 825 | 100% |
| Corticostriatal Tract R Anterior | 176 | 179 | 98.3% |
| Superior Longitudinal Fasciculus R 2 | 114 | 264 | 43.2% |
| Thalamic Radiation R Anterior | 94 | 94 | 100% |
| Corpus Callosum Forceps Minor | 88 | 100 | 88% |
| Reticular Tract R | 76 | 86 | 88.4% |
| Uncinate Fasciculus R | 52 | 52 | 100% |
| Corpus Callosum Body | 43 | 400 | 10.8% |
| Corticostriatal Tract R Superior | 37 | 224 | 16.5% |
| Cingulum R Parolfactory | 33 | 37 | 89.2% |
| Arcuate Fasciculus R | 33 | 137 | 24.1% |
| Anterior Commissure Frontal | 32 | 32 | 100% |
| Non Decussating Dentatorubrothalamic Tract R | 30 | 146 | 20.5% |
| Thalamic Radiation R Superior | 29 | 171 | 17% |
| Cingulum R Frontal Parietal | 28 | 121 | 23.1% |

##### E2B. Bundles by calculated-dose category — ≥120 Gy

| **Bundle** | **Streamlines in category** | **Streamlines in bundle** | **% in category** | **Peak dose (Gy)** |
| --- | --- | --- | --- | --- |
| Inferior Fronto Occipital Fasciculus R | 674 | 825 | 81.7% | 1231.9 |
| Corticostriatal Tract R Anterior | 147 | 179 | 82.1% | 1623.4 |
| Reticular Tract R | 74 | 86 | 86% | 372.1 |
| Thalamic Radiation R Anterior | 68 | 94 | 72.3% | 275.2 |
| Uncinate Fasciculus R | 37 | 52 | 71.2% | 197.8 |
| Corpus Callosum Forceps Minor | 35 | 100 | 35% | 614.1 |
| Corticopontine Tract R Frontal | 27 | 82 | 32.9% | 897.1 |
| Arcuate Fasciculus R | 17 | 137 | 12.4% | 303.3 |
| Corticostriatal Tract R Superior | 16 | 224 | 7.1% | 1577.3 |
| Anterior Commissure Frontal | 14 | 32 | 43.8% | 262.7 |
| Superior Longitudinal Fasciculus R 2 | 12 | 264 | 4.5% | 161.1 |
| Extreme Capsule R | 9 | 63 | 14.3% | 200.2 |

##### E2C. Bundles by calculated-dose category — 60 to <120 Gy

| **Bundle** | **Streamlines in category** | **Streamlines in bundle** | **% in category** | **Peak dose (Gy)** |
| --- | --- | --- | --- | --- |
| Superior Longitudinal Fasciculus R 2 | 42 | 264 | 15.9% | 161.1 |
| Ansa Lenticularis R | 42 | 59 | 71.2% | 76.3 |
| Arcuate Fasciculus R | 40 | 137 | 29.2% | 303.3 |
| Ansa Subthalamica R | 26 | 62 | 41.9% | 63.8 |
| Corticostriatal Tract R Superior | 23 | 224 | 10.3% | 1577.3 |
| Fasciculus Subthalamicus R | 20 | 70 | 28.6% | 76.8 |
| Cingulum R Frontal Parietal | 17 | 121 | 14% | 111.5 |
| Fasciculus Lenticularis R | 16 | 26 | 61.5% | 63.3 |
| Thalamic Radiation R Anterior | 14 | 94 | 14.9% | 275.2 |
| Corticostriatal Tract R Anterior | 14 | 179 | 7.8% | 1623.4 |
| Corpus Callosum Body | 13 | 400 | 3.2% | 256.4 |
| Anterior Commissure Temporal | 12 | 54 | 22.2% | 100.7 |

##### E2D. Cortical parcels by maximum sampled dose (HCP-MMP)

| **Parcel** | **Anatomical location** | **Max dose (Gy)** |
| --- | --- | --- |
| R_11l | Orbitofrontal | 1590.6 |
| R_47l | Orbitofrontal | 1558.4 |
| R_MI | Insula Proper | 1534.3 |
| R_FOP4 | Superior Opercula | 1019.5 |
| R_pOFC | Orbitofrontal | 892.4 |
| R_AAIC | Insula Proper | 800.2 |

*Maximum calculated dose sampled within each HCP-MMP parcel; ranking is within-case only.*

#### Table E3. Patient 3 — diffuse right fronto-temporo-parietal glioblastoma

*Diffuse right fronto-temporo-parietal glioblastoma, preoperative virtual plan.*

##### E3A. Bundles intersecting the projected resection

| **Bundle** | **Intersecting streamlines** | **Streamlines in bundle** | **% intersecting** |
| --- | --- | --- | --- |
| Inferior Fronto Occipital Fasciculus R | 199 | 825 | 24.1% |
| Uncinate Fasciculus R | 36 | 52 | 69.2% |
| Anterior Commissure Frontal | 29 | 32 | 90.6% |
| Corpus Callosum Forceps Minor | 20 | 100 | 20% |
| Corticostriatal Tract R Anterior | 10 | 179 | 5.6% |

##### E3B. Bundles by calculated-dose category — ≥120 Gy

| **Bundle** | **Streamlines in category** | **Streamlines in bundle** | **% in category** | **Peak dose (Gy)** |
| --- | --- | --- | --- | --- |
| Inferior Fronto Occipital Fasciculus R | 333 | 825 | 40.4% | 1920.6 |
| Uncinate Fasciculus R | 47 | 52 | 90.4% | 1151.3 |
| Anterior Commissure Frontal | 32 | 32 | 100% | 1470.3 |
| Corticostriatal Tract R Anterior | 26 | 179 | 14.5% | 1150.5 |
| Corpus Callosum Forceps Minor | 26 | 100 | 26% | 842.3 |
| Inferior Fronto Occipital Fasciculus L | 5 | 447 | 1.1% | 180.6 |
| Cingulum R Frontal Parietal | 3 | 121 | 2.5% | 177.5 |
| Superior Longitudinal Fasciculus R 2 | 1 | 264 | 0.4% | 124.9 |
| Thalamic Radiation R Anterior | 1 | 94 | 1.1% | 139 |

##### E3C. Bundles by calculated-dose category — 60 to <120 Gy

| **Bundle** | **Streamlines in category** | **Streamlines in bundle** | **% in category** | **Peak dose (Gy)** |
| --- | --- | --- | --- | --- |
| Inferior Fronto Occipital Fasciculus R | 73 | 825 | 8.8% | 1920.6 |
| Inferior Fronto Occipital Fasciculus L | 14 | 447 | 3.1% | 180.6 |
| Corticostriatal Tract R Anterior | 8 | 179 | 4.5% | 1150.5 |
| Thalamic Radiation R Anterior | 5 | 94 | 5.3% | 139 |
| Superior Longitudinal Fasciculus R 2 | 5 | 264 | 1.9% | 124.9 |
| Cingulum R Frontal Parietal | 4 | 121 | 3.3% | 177.5 |
| Reticular Tract R | 4 | 86 | 4.7% | 68.4 |
| Corpus Callosum Forceps Minor | 4 | 100 | 4% | 842.3 |
| Corticostriatal Tract L Anterior | 3 | 209 | 1.4% | 108.2 |
| Reticular Tract L | 1 | 138 | 0.7% | 72 |
| Thalamic Radiation L Anterior | 1 | 121 | 0.8% | 112.7 |
| Uncinate Fasciculus R | 1 | 52 | 1.9% | 1151.3 |

##### E3D. Cortical parcels by maximum sampled dose (HCP-MMP)

| **Parcel** | **Anatomical location** | **Max dose (Gy)** |
| --- | --- | --- |
| R_11l | Orbitofrontal | 3483.7 |
| R_10r | Subgenual Cingulate | 1598 |
| R_a10p | Frontopolar | 906.4 |
| R_10pp | Frontopolar | 783.3 |
| R_OFC | Orbitofrontal | 539 |
| R_p10p | Frontopolar | 523.8 |

*Maximum calculated dose sampled within each HCP-MMP parcel; ranking is within-case only.*

#### Table E4. Case and imaging summary

| **Patient** | **Anatomical description** | **Preoperative target definition** | **Postoperative MRI** | **Clinical dose comparator** |
| --- | --- | --- | --- | --- |
| 1 | Left parieto-temporal glioblastoma | Enhancing T1-weighted MRI | Available | Available |
| 2 | Right frontal glioblastoma | Enhancing T1-weighted MRI | Available | Unavailable |
| 3 | Diffuse right fronto-temporo-parietal glioblastoma | T1 enhancement and FLAIR-defined nonenhancing frontal component | Available | Unavailable |

Note. Case descriptions reflect the imaging available for this proof-of-concept analysis.

#### Table E5. Detailed Patient 1 validation metrics

| **Metric** | **80 Gy** | **60 Gy** |
| --- | --- | --- |
| Volumetric agreement |  |  |
| Clinical volume (mL) | 20.89 | 31.63 |
| Calculated volume (mL) | 21.33 | 31.89 |
| Shared volume (mL) | 15.60 | 25.47 |
| Calculated / clinical volume | 1.021 | 1.008 |
| Dice similarity | 0.7393 | 0.8021 |
| Jaccard index | 0.5864 | 0.6695 |
| Sensitivity | 74.7% | 80.5% |
| Positive predictive value | 73.2% | 79.9% |
| Centroid offset (mm) | 3.48 | 3.56 |
| Centroid offset, in-plane component (mm) | 3.42 | 3.49 |
| Normative HCP-1065 streamline-set and bundle agreement |  |  |
| Clinical-volume streamlines | 421 | 541 |
| Calculated-volume streamlines | 558 | 920 |
| Shared streamlines | 415 | 541 |
| Clinical streamlines captured | 98.6% | 100.0% |
| Calculated-only streamlines | 143 | 379 |
| Streamline-set Dice | 0.8478 | 0.7406 |
| Streamline-set Jaccard | 0.7358 | 0.5880 |
| Calculated / clinical streamline count | 1.325 | 1.701 |
| Bundle Spearman ρ (bundles compared) | 0.9604 (14) | 0.7887 (17) |
| Bundle Pearson r | 0.9361 | 0.9305 |
| Shared bundles among top 10 | 9/10 | 9/10 |

Note. The calculation used 3.03 U per seed. Sensitivity is the fraction of the clinical volume recovered; positive predictive value is the fraction of calculated volume inside the clinical volume. Streamline counting and bundle eligibility follow Sections 2.6–2.7.

### Supplementary Figures

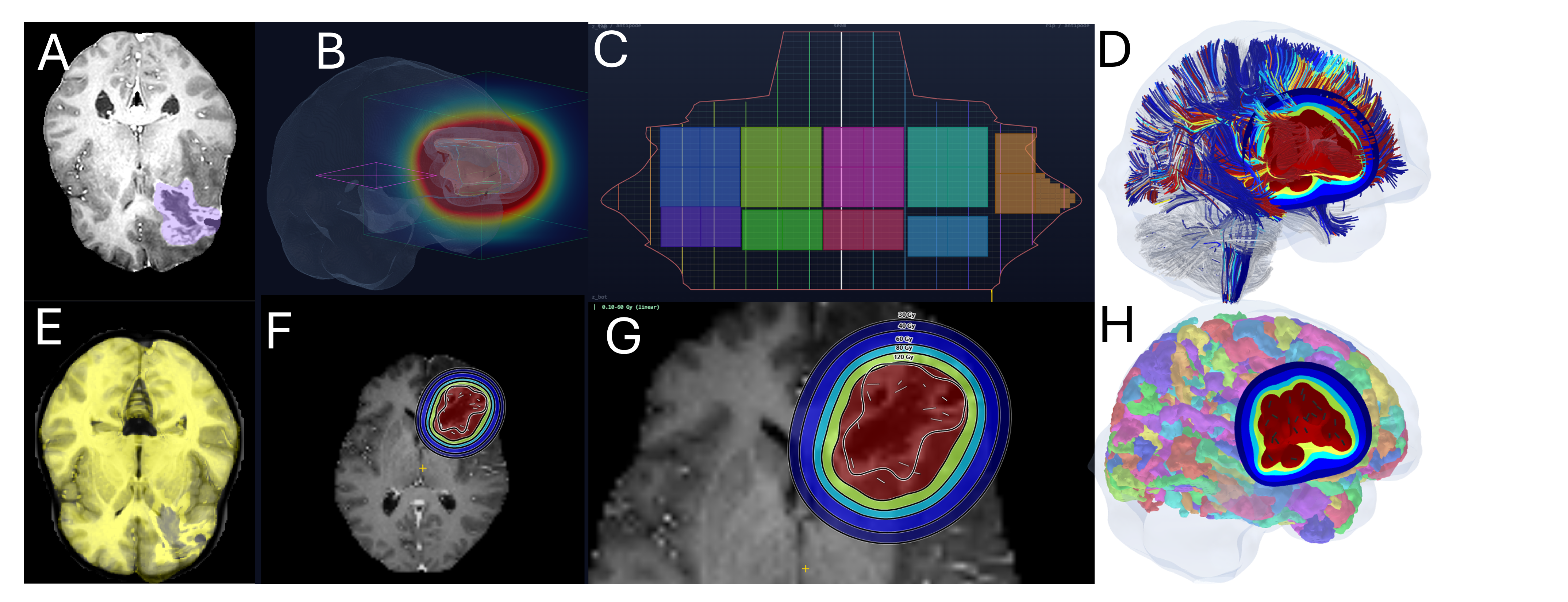

Supplementary Figure E1. Patient 2 preoperative virtual planning and modeled neuroanatomical exposure. (A) Axial contrast-enhanced T1-weighted MRI with the observed right frontal target in violet. (B) Patient-derived representation of the projected resection surface with the virtual tile arrangement and calculated dose field. (C) Opened planning map with colored virtual tile footprints. (D) Blended patient-to-MNI registration overlay. (E) Axial overview of the 30-, 40-, 60-, 80-, and 120-Gy calculated dose bands on the patient MRI. (F) Magnified axial dose view showing virtual seed positions. (G) HCP-MMP cortical parcels with calculated dose bands overlaid. (H) Normative HCP-1065 tractogram with atlas streamlines colored by their maximum sampled dose. All panels use the projected preoperative surface representation.

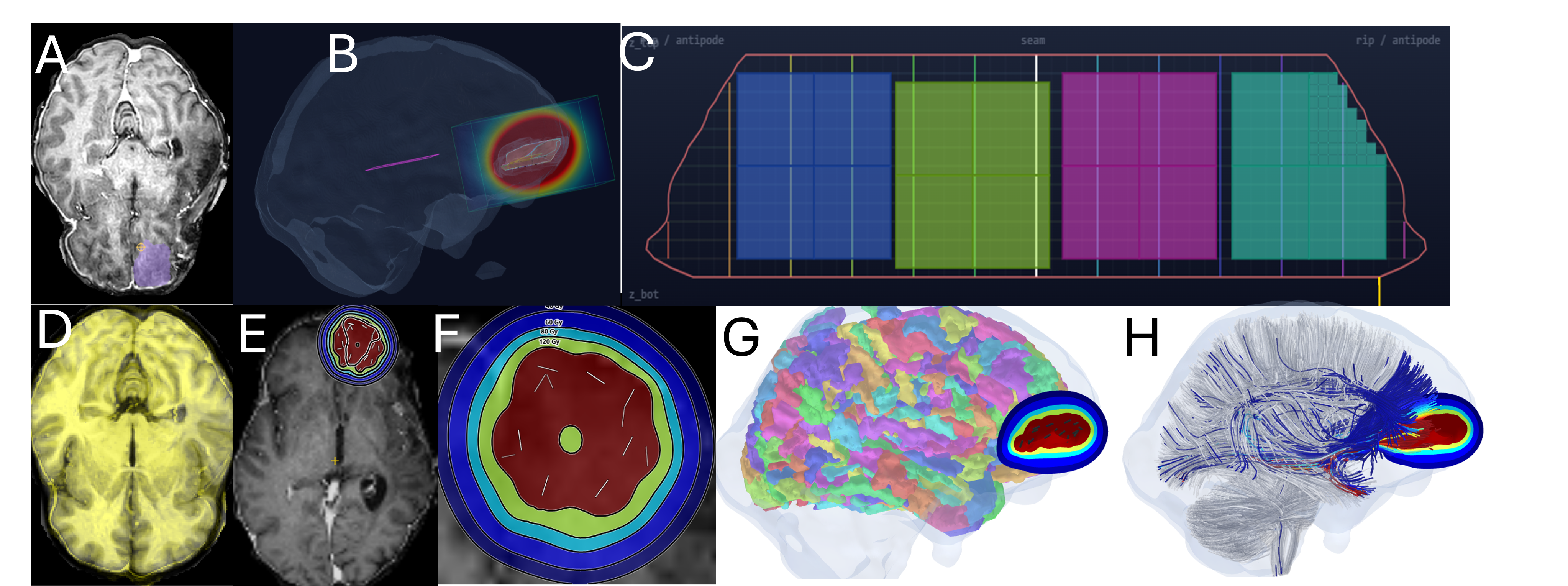

Supplementary Figure E2. Patient 3 preoperative virtual planning and modeled neuroanatomical exposure. (A) Axial preoperative MRI with the observed right fronto-temporo-parietal target in violet. (B) Patient-derived representation of the projected resection surface with the virtual tile arrangement and calculated dose field. (C) Opened planning map with colored virtual tile footprints. (D) Blended patient-to-MNI registration overlay. (E) Axial overview of the 30-, 40-, 60-, 80-, and 120-Gy calculated dose bands on the patient MRI. (F) Magnified axial dose view showing virtual seed positions. (G) HCP-MMP cortical parcels with calculated dose bands overlaid. (H) Normative HCP-1065 tractogram with atlas streamlines colored by their maximum sampled dose. All panels use the projected preoperative surface representation.
